# Assessing COVID prevention strategies to permit the safe opening of college campuses in fall 2021

**DOI:** 10.1101/2021.07.19.21260522

**Authors:** A. David Paltiel, Jason L. Schwartz

## Abstract

**Background:** Effective vaccines, improved testing technologies, and declines in COVID-19 incidence prompt an examination of the choices available to college administrators to safely resume in-person campus activities in fall 2021.

**Objective:** To develop a decision support tool that assists college administrators in designing and evaluating customized COVID vaccination, screening, and prevention plans.

**Design:** Decision analysis linked to a compartmental epidemic model, quantifying the interaction of policy instruments (e.g., vaccination promotion, asymptomatic testing, physical distancing, and other non-pharmaceutical interventions), institutional priorities (e.g., risk tolerance, desire to resume activities), and assumptions about vaccine performance and background epidemic severity.

**Participants:** Hypothetical cohort of 5000 individuals (students, faculty, and staff) living and working in the close environs of a residential college campus.

**Main Outcome(s) and Measure(s):** Cumulative infections over a 120-day semester.

**Results:** Under Base Case assumptions, if 90% coverage with an 85%-effective vaccine can be attained, the model finds that campus activities can be fully resumed while holding cumulative cases below 5% of the population without the need for routine, asymptomatic testing. With 50% population coverage using such a vaccine, a similar “return to normalcy” would require daily asymptomatic testing of unvaccinated individuals. The effectiveness of vaccination in reducing susceptibility to infection is a critical uncertainty.

**Conclusions & Relevance:** Vaccination coverage is the most powerful tool available to college administrators to achieve a safe return to pre-pandemic operations this fall. Given the breadth of potential outcomes in the face of uncontrollable and uncertain factors, even colleges that achieve high vaccination coverage should be prepared to reinstitute testing and distancing policies on short notice.

## INTRODUCTION

Institutions of higher education (IHEs) throughout the United States are once again confronting the challenges posed by SARS-CoV-2 in their planning for safe operations during the approaching academic year. As in 2020, the characteristic features of campus life – communal living arrangements with shared dining, sleeping, and bathing spaces; classrooms, performance spaces, and athletic venues of varying size and density; and a population of young adults eager to socialize – raise concern about superspreading and the safety of surrounding communities.^1^

This year, however, IHE administrators are presented with a dramatically more favorable context upon which to act than they encountered in summer 2020. Chief among these differences is the widespread availability of multiple highly effective vaccines. Vaccines in use in the United States have been shown to dramatically reduce the incidence of symptomatic disease.^2^ Additional evidence demonstrates significant reductions in viral transmission and infections, notably asymptomatic infections.^3^ Breakthrough infections among vaccinated persons are exceedingly rare and generally minor in severity.^4^

In addition to vaccines, testing for SARS-CoV-2—a central element of the early response to COVID-19—remains a potent tool in the public health arsenal. The availability and design of testing has expanded considerably throughout the pandemic.^5^ Routine, asymptomatic screening can now be performed effectively and conveniently at IHEs at marginal costs as low as $2.50 per test.^6^

The experience of the last year has likewise demonstrated the effectiveness of non-pharmaceutical interventions (NPIs) in reducing the spread of the virus.^7^ IHEs that adhered assiduously to a combination of high-cadence testing, de-densification, masking, hand-washing, and social distancing appear to have virtually eliminated the transmission of infection on their campuses during the 2020-21 academic year.^8^

These observations suggest that the question facing IHE officials is not whether there exists a safe approach to bringing students onto campus this fall. Rather, their challenge is how best to design a portfolio of strategies, a combination of vaccination policies, virologic monitoring, and NPIs, that will strike what they consider an appropriate balance between adequate outbreak control—amid what remains a serious and uncertain threat in the months ahead—and a satisfactory restoration of the residential college experience.

The purpose of this report is to provide evidence and structure to address that challenge. We offer a quantitative framework that supports exploration of policy approaches and evaluates the feasibility and mutual compatibility of different combinations of strategies and performance targets. Because background conditions and risk tolerance will vary among IHEs and their epidemiological settings, we have also developed a publicly available <u>spreadsheet implementation</u> of our model-based analysis. This companion tool permits IHE decision-makers not only to reproduce the results reported here but also to design a customized portfolio of strategies that reflects their institution’s specific priorities and circumstances.

## METHODS

### Analytic overview

We developed a decision framework to inform the design of a college reopening strategy for the fall 2021 semester. Choices available to decision-makers in this framework include: anticipated vaccination coverage among the campus community (the product of the policies ranging from education to incentives to requirements with varying enforcement or availability of exemptions); virologic monitoring (testing) strategies with varying frequencies of testing and target populations; and the continued use of NPIs (e.g., distancing, masking), all considered in the context of a goal of returning to normal (i.e., pre-COVID-19) operations and activities, in whole or in part. We linked this framework to a simple, state-transition model of SARS-CoV-2 transmission on campus.^1^ This permitted us to capture the dynamic interaction of alterative policy interventions and to measure their combined impact on cumulative infections over a 120-day semester.

### Policy instruments and preferences

Motivating our approach is the contention that those responsible for setting policies for IHEs—e.g., school administrators, state-appointed overseers, government education officials, and/or governors or other elected officials—express their beliefs, priorities, and preferences regarding campus safety and operations via the choices they make on the following four dimensions of decision-making:

#### 1) Target vaccination coverage level

Rates of vaccination on college campuses this fall will vary widely, mirroring the significant differences in both vaccination coverage across U.S. regions and the intensity with which IHE officials will encourage vaccination on campus. To date, more than 500 IHEs are requiring students to be vaccinated,^9^ with varying degrees of enforcement and availability of exemptions for non-medical (in addition to medical) reasons. Some of those institutions have extended that requirement to faculty and staff. The great majority of the nation’s ∼4,000 IHEs are not, at present, requiring vaccination, a possible response to political pressures or mandates from elected officials, the current regulatory status of the authorized vaccines, the potential for declines in enrollments, or other considerations.^10–11^

Colleges that elect to do nothing to actively encourage vaccination will nevertheless benefit from ongoing national vaccination efforts and can plausibly anticipate that 50% of their campus population will be vaccinated.^12^ (This figure reflects the comparatively lower rate of vaccination among the college-age population to date.) Colleges that take a more active approach may elect to do so with varying degrees of aggressiveness. For purposes of illustration, we have chosen 70% as a mid-range value, representing schools that actively encourage or create incentives for vaccination but do not require it. We adopt 90% as our upper-bound value, one that should be attainable by colleges that vigorously ensure compliance with an explicit vaccination requirement. Vaccination rates closer to 95% or greater may be achieved depending on the availability and ease of obtaining non-medical exemptions and the level of monitoring and enforcement.

#### 2) Testing frequency and targeting

Routine, asymptomatic screening for SARS-CoV-2 infection, coupled with contact tracing and isolation following positive results, were considered a cornerstone for containment and control of campus outbreaks in the fall of 2020.^13^ This year, colleges may choose from a broader variety of more convenient, less costly testing alternatives. Current published guidance offers conflicting views on the need for asymptomatic screening.^6, 14, 15^ In situations where screening is recommended, guidelines differ on both the frequency and targeting strategy. Some IHEs may elect to target screening activities solely toward unvaccinated individuals; others may test all members of the campus community, regardless of vaccination status; still others may retire their asymptomatic testing programs and test only symptomatic individuals. In the illustrative examples that follow, we consider both targeted and untargeted testing, at rates ranging from once every 2 weeks to daily. We also consider the option of restricting testing only to persons with symptoms.

#### 3) The tradeoff between “safety” and “normalcy”

The balance between safety and the return to normalcy is a quantifiable measure of institutional priorities and preferences. It represents a choice that can be struck at any point on the continuum that runs between a total lockdown at one extreme and an unrestricted return to pre-COVID activities at the other extreme. IHE administrators control that choice and reveal their preferences by where they elect to place their school on that safety-versus-normalcy continuum. This continuum is not just a conceptual abstraction; one may quantify choice of location on that continuum using the effective, on-campus reproduction number (Rt), taking into account a college’s adoption of NPIs against the background epidemiology of the pandemic in the surrounding community.^16^

In fall 2020, the choice on some of the nation’s campuses tilted heavily toward safety—with little-to-no in-person instruction; minimal athletics, performances, or other extracurriculars; de-densified dormitories; large-scale asymptomatic screening programs, distancing, masking, and other measures. The Rt on these campuses may have been close to zero. On other campuses, attention to NPIs was less aggressive and the on-campus Rt likely exceeded its off-campus analog. Statewide estimates of Rt in the US currently range from roughly 0.5 to 1.5.^17^ However, recent reports suggest that new viral variants may have reproductive numbers as high as 7.^18^ In the illustrative examples that follow, we adopt a conservative baseline value of 3, reflecting the desire of many college decision-makers to return to the intimacy of pre-COVID, residential campus activities without physical distancing but also accounting for the threat of new variants.^6^

#### 4) Tolerance for campus infections

Eliminating all viral transmission is incompatible with any resumption of on-campus activities this fall. A more credible expectation is that the cumulative number or rate of infections on campus might be held below some ceiling that has been deemed “tolerable.” Such a threshold would serve as a benchmark to evaluate the success of a given strategy in meeting its safety objectives. College administrators might balk at the prospect of specifying an explicit threshold. However, the degree to which “acceptable campus infections” constitutes a choice—one that reflects decision-makers’ preferences and institutional priorities— can be inferred from the widely divergent responses to outbreaks observed during the 2020-21 academic year. The University of North Carolina at Chapel Hill, for example, abandoned an attempt to hold most fall classes in person in the very first week of the semester, moving online after fewer than 200 of its ∼30,000 students tested positive for the coronavirus.^19^ By contrast, the University of Wisconsin-Madison (∼31,600 students) remained open for the entire year, even after 911 students and staff tested positive in a single week in September and more than7700 cases were detected over the academic year.^20^ For purposes of illustration, we use a ceiling of 5%, noting that this figure includes both asymptomatic and symptomatic cases.

In articulating these four dimensions of decision-making, we make no value judgments. Rather, we treat each dimension as an independent choice that reveals useful information about decision-makers’ preferences, priorities, and constraints. Our aim is to assemble these choices and to report on their mutual compatibility, given current knowledge about the virus, its epidemiology, and the impact of vaccines and NPIs. If a set of choices is not feasible, the epidemic model can point to the additional concessions or responses that might be required.

### Analytic model

We updated the simple compartmental epidemic model first developed for our July 2020 analysis of campus screening options.^1^ A full description of the model and input data assumptions is provided in the Technical Appendix. This model captures many of the essential features of the situation facing college decision-makers: the epidemiology of SARS-CoV-2; the natural history of COVID-19 illness; and the availability of testing technologies to detect, isolate, and contain the presence of SARS-CoV-2 in a residential college setting (**Appendix Figure S1**). The updated model adds functionality to account for the presence of individuals who have achieved antibody protection, either as the result of prior infection with SARS-CoV-2 or via vaccination. These individuals are assumed to be at reduced risk of acquiring and transmitting infection. They are further assumed to progress to symptomatic and advanced COVID-19 at lower rates, if and when breakthrough infections do occur. Model input data were obtained from a variety of published sources (**Table 1** and Appendix).^12, 21–32^

**Table 1.**
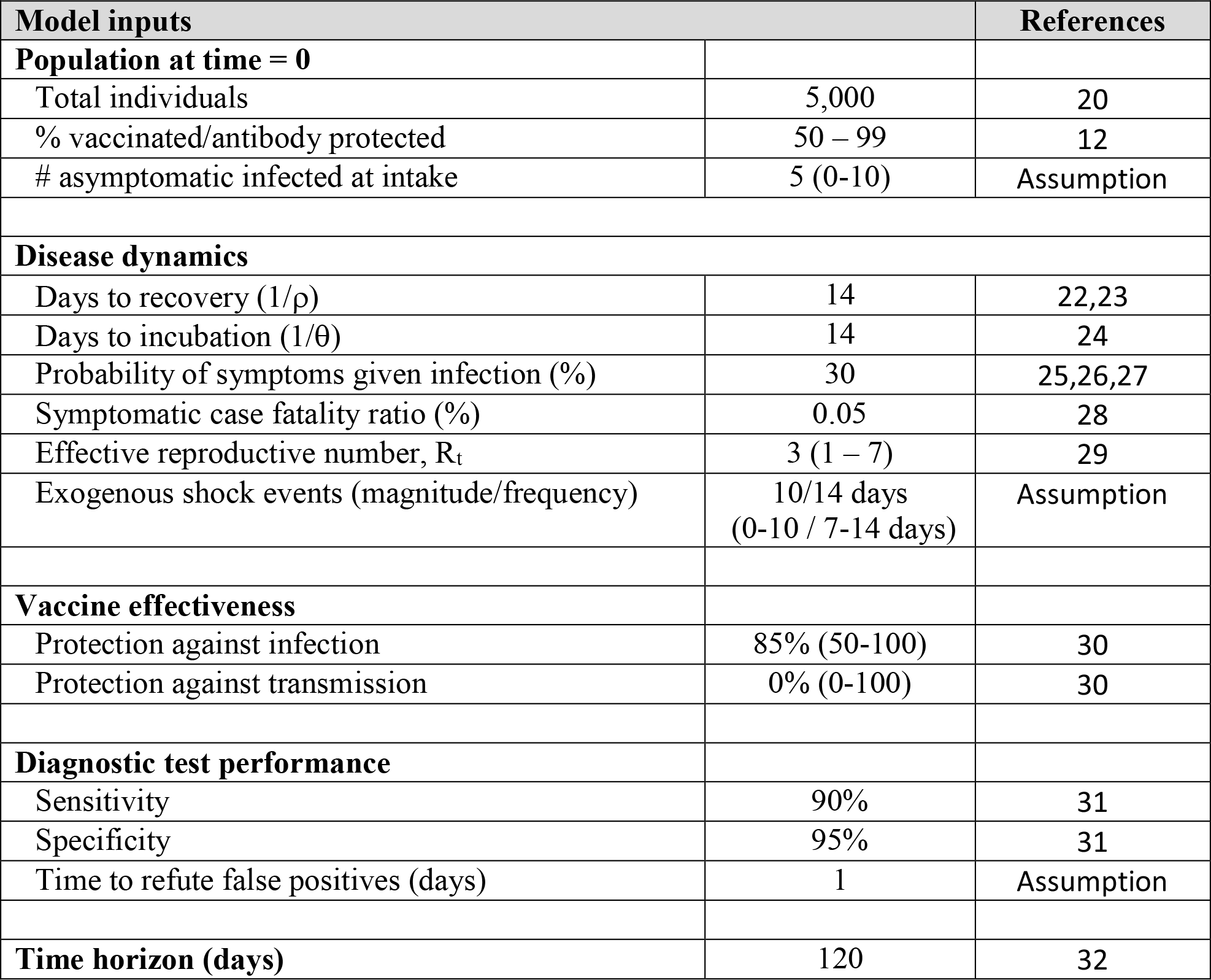
Model input parameters.

### Sensitivity analysis

Beyond the four dimensions of decision-making described above, there are a host of factors and uncertainties over which college administrators have no influence. To help decision-makers understand how the consequences of their choices might vary in the face of variation in these uncontrollable factors, we conducted sensitivity analysis on vaccine effectiveness (range 75%-90%), the frequency and magnitude of exogenous “imported” infections, and the presence of undetected, asymptomatic individuals on campus at the start of the semester. Interested readers may conduct additional explorations using the publicly accessible implementation of the model at [LINK].

This analysis adheres to the Consolidated Health Economic Evaluation Reporting Standards (CHEERS) reporting guideline, where applicable.^33^

## RESULTS

**Figure 1** reports the findings of the model under baseline input data assumptions, assuming 50% coverage with an 85%-effective vaccine. A “return to normalcy” (Rt > 3, as defined previously) while holding cumulative infections below 5% of the student population (green box) can only be achieved if routine, asymptomatic testing of unvaccinated individuals occurs daily (purple curve) or perhaps every 3 days (orange line). In the absence of any testing, infections could still be held below 5%, but Rt would need to fall below 1.2, conditions consistent with the resumption of aggressive distancing and other prevention policies. Without either testing or distancing policies, a return to pre-COVID campus life and activities could result in the infection of virtually all unvaccinated members of the population before the end of the semester (extending the blue curve beyond the boundaries of the figure).

**Figure 1.**
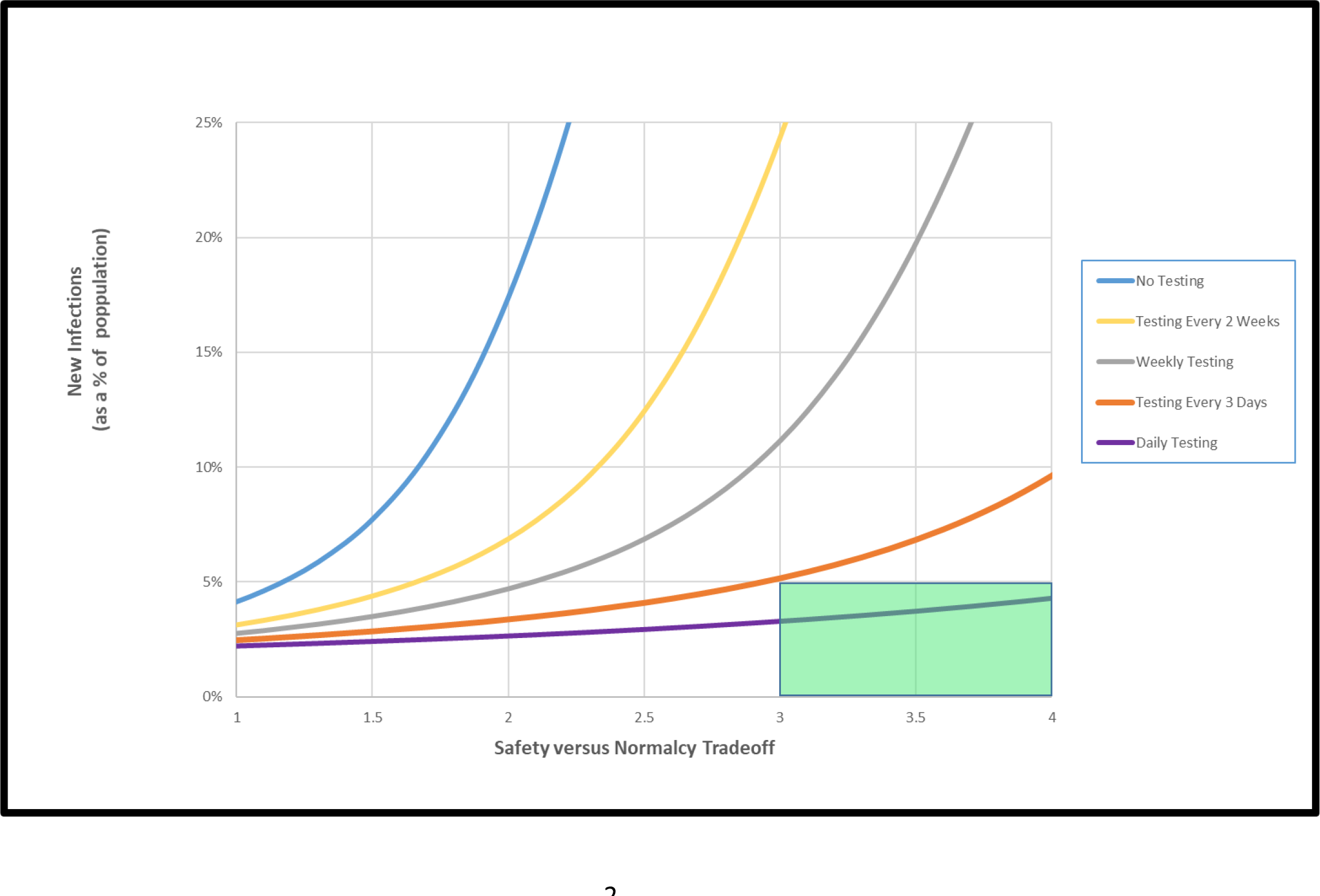
Cumulative infections with 50% vaccine coverage, 85% vaccine effectiveness, and asymptomatic testing targeted to unvaccinated population. The figure reports the findings of the model under baseline input data assumptions, assuming 50% coverage with an 85%-effective vaccine. The green box denotes the circumstances under which the “return to normalcy” (Rt > 3, as defined previously) can be achieved while holding cumulative infections below 5% of the campus population. The curves denote the expected cumulative infections under alternative frequencies of routine, asymptomatic testing of unvaccinated individuals (ranging from daily testing to no asymptomatic testing).

Raising vaccination coverage to 70% produces small improvements in the model’s predicted outcomes (**Figure 2**). Routine, asymptomatic testing of unvaccinated persons every 3 days (and possibly every week) might permit the return to normalcy while holding cumulative case rates below 5% of the student population (green box).

**Figure 2.**
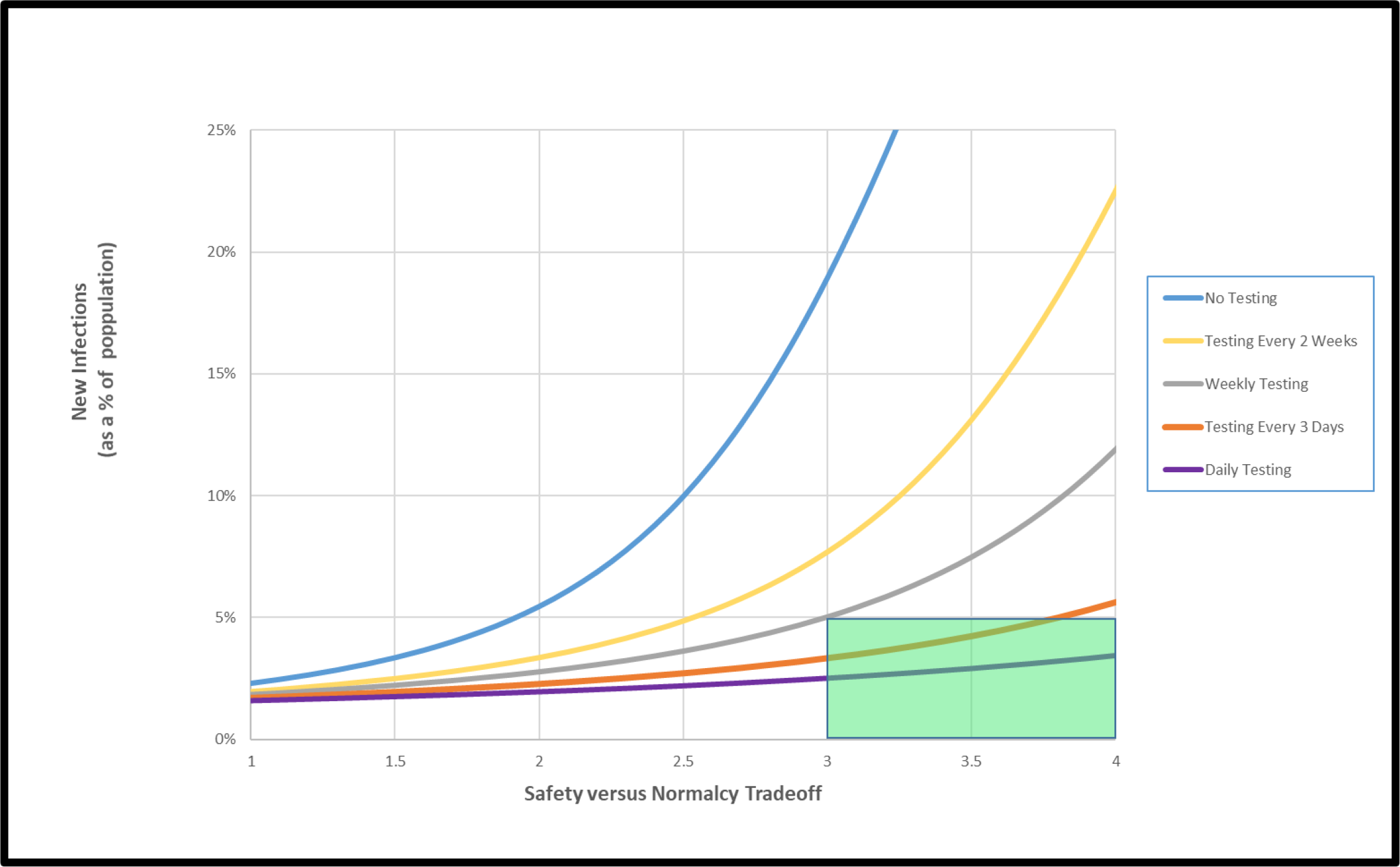
Cumulative infections with 70% vaccine coverage, 85% vaccine effectiveness, and asymptomatic testing targeted to unvaccinated population. The figure reports the findings of the model under baseline input data assumptions, assuming 70% coverage with an 85%-effective vaccine. The green box denotes the circumstances under which the “return to normalcy” (Rt > 3, as defined previously) can be achieved while holding cumulative infections below 5% of the campus population. The curves denote the expected cumulative infections under alternative frequencies of routine, asymptomatic testing of unvaccinated individuals (ranging from daily testing to no asymptomatic testing).

Cumulative infections could be held below 10% either by testing every 2 weeks or by implementing moderate distancing efforts (Rt = 2.5).

The model suggests that the need for any asymptomatic testing would be eliminated at colleges that can achieve vaccination coverage levels of 90% (**Figure 3**).

**Figure 3.**
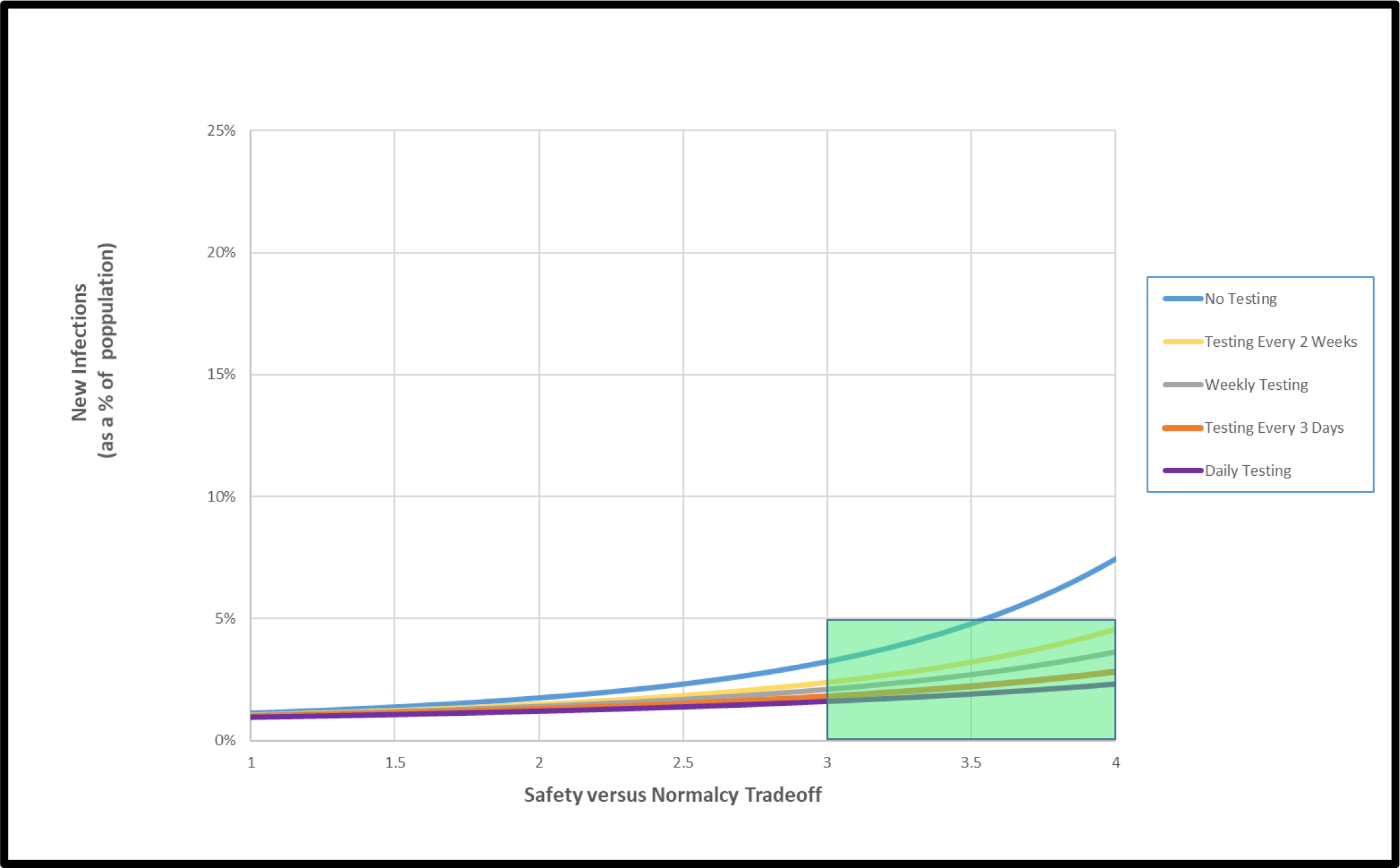
Cumulative infections with 90% vaccine coverage, 85% vaccine effectiveness, and asymptomatic testing targeted to unvaccinated population. The figure reports the findings of the model under baseline input data assumptions, assuming 90% coverage with an 85%-effective vaccine. The green box denotes the circumstances under which the “return to normalcy” (Rt > 3, as defined previously) can be achieved while holding cumulative infections below 5% of the campus population. The curves denote the expected cumulative infections under alternative frequencies of routine, asymptomatic testing of unvaccinated individuals (ranging from daily testing to no asymptomatic testing).

### Sensitivity analysis

#### Vaccine effectiveness

In general, less optimistic assumptions regarding vaccine preventive effectiveness necessitate greater attention to screening and distancing options. Even with high vaccination coverage (90%), the model suggests that an 80%-effective vaccine would need to be supplemented with infrequent (i.e., every 2 weeks) testing or with limited NPIs (Rt = 2.7) to hold cumulative case rates below 5% (**Supplemental Figure S1A)**. An 80%-effective vaccine would necessitate daily testing at any college where the vaccination coverage rate was below 70% (**Supplemental Figure S1B**).

#### Universal testing

Expanding the screening effort to all members of the campus community, regardless of vaccination status, invariably reduces the cumulative number of infections. At colleges with low (50%) vaccination coverage, this could have a small but noticeable impact on policy: it might be possible to reduce the frequency of testing from daily to once every 3 days (**Supplemental Figure S2A**). On campuses with high rates of vaccination (90%), expanding screening to the entire population would carry little incremental benefit (**Supplemental Figure S2B**).

## DISCUSSION

As colleges prepare for the fall 2021 semester, several conclusions with implications for their planning emerge from this modeling exercise, including the paradigmatic scenarios presented above and the countless alternative designs that can be examined using the publicly available <u>companion tool</u>.

First, one size does not fit all. No single, “optimal” plan for college reopening captures the breadth of institutional circumstances and priorities on campuses nationwide. The curves presented in the Results section are highly malleable and dependent on idiosyncratic choices that fall within the control of decision-makers. These choices include: the degree to which they seek a “normal” or “close-to-normal” campus experience through the relaxation of NPIs; the efforts they undertake to promote or ensure very high vaccination rates, including the potential use of requirements; and the number of cases they deem to be within acceptable levels. IHEs can and should therefore tailor their public health strategies to their particular settings, conditions, and concerns, recognizing the trade-offs inherent in any policy approach. The web-based companion app is available to support such an exercise.

Second, develop an adaptive contingency plan. Just as there is considerable uncertainty with respect to the trajectory of the COVID-19 pandemic in the months ahead, so too is there uncertainty regarding the suitability of any static approach intended to facilitate the safe return to on-campus college life over a months-long time horizon. The curves presented in the Results section are highly sensitive to the uncertainties in the underlying data. Plausible variation in factors beyond the control of decision-makers – the preventive effectiveness of the vaccine or the emergence of a more transmissible viral variant, to cite just two possibilities – can produce greatly divergent outcomes. Given that some of these uncertain factors can change rapidly, even IHEs that enter fall 2021 in an apparent favorable position should remain vigilant and prepared to rapidly alter their approaches—to vaccination policies; distancing, making, and other NPIs; testing, or a combination thereof—if conditions warrant.

Third, maximize vaccine coverage. Of the many factors within the control of IHE decision-makers planning for fall 2021, the level of vaccination in the campus community is likely to be the single most powerful determinant of safety, as measured through the number of cases occurring among students, faculty, and staff. IHEs with high vaccination rates—particularly those exceeding 90%—can insulate themselves against most of the threats that will otherwise prompt widespread or frequent testing and a return to intensive NPIs. Vaccination also offers the most reliable envelope of protection in the face of potential worsening public health conditions leading to increased Rt.

Fourth, monitor emerging data on vaccine effectiveness. Of the many factors that lie beyond the control of decision-makers planning for fall 2021, none is as worrisome as a potential decline in vaccine effectiveness. Students, faculty, and staff will arrive on campus having been vaccinated using any number of vaccines—one of the three authorized by the U.S. Food and Drug Administration or one of the several other vaccines in use internationally. While the U.S.-authorized vaccines all show high levels of protection against severe outcomes, their overall effectiveness varies. Less is known about the effectiveness of international vaccines authorized by the World Health Organization, particularly the protection they provide against variants. While the current vaccines have largely remained resilient against the challenges presented by current variants, even modest reductions in overall vaccine effectiveness in the months ahead could significantly alter the degree to which a campus reopening plan is adequate to provide the level of community protection sought by decision-makers. Careful attention by IHE officials to emerging evidence regarding vaccine effectiveness and related guidance from local, state, and health officials will be essential to a sustainable fall 2021 semester on-campus.

Fifth, carefully consider the targeting of testing activities. Decision-makers have the option not only to institute a program of routine asymptomatic testing for SARS-CoV-2 but also to determine whether that program should be applied to all members of the campus community or only those who remain unvaccinated. Our exploratory investigations suggest that the benefits of extending testing to the broader population may be modest unless campus vaccination rates are very low (i.e., ∼50%) or the vaccine is found to be much less effective than current evidence has shown. In addition to those limited benefits of widespread testing, such programs are financially costly to colleges and increase the risk of false positives and resulting disruptions and confusion. IHEs that limit required, routine testing only to unvaccinated members of their community also gain the opportunity for that policy to serve as an incentive for individuals to be vaccinated.

The simple model underlying this analysis has notable limitations. We assumed a homogenous population encompassing all persons—students, faculty, and staff—living and working in close, regular physical proximity to one another in a campus setting. We did not explicitly take into account non-random mixing patterns, age-dependent transmission, or any of the other evident differences between students and university employees, although non-student members of the college community include a higher proportion of older, more medically vulnerable individuals. Also, we do not distinguish between students living in dormitories and those living in off-campus settings. These are important distinctions deserving further exploration, but they are beyond the scope of the present analysis.

Recent guidance documents for IHEs from the Centers for Disease Control and Prevention, the American College Health Association, the Massachusetts Higher Education Testing Group, and other organizations have been released to support decision-making by IHE officials preparing for a safe reopening for the fall 2021 semester while restoring at least a significant degree of normalcy to the on-campus experience.^6, 13, 14^ Our work aims to inform and enhance that work, providing a tool that illuminates the complex interplay of choices available to them as they consider their institutional circumstances, priorities, and needs in the face of an ongoing and uncertain public health threat.

## Data Availability

All data used in this analysis were obtained from published, publicly available sources, which we have cited in the manuscript references.

## ACKNOWLEDGMENT

**Funding Sources:** This work was supported by the National Institute on Drug Abuse of the National Institutes of Health via award R37 DA015612. The content is solely the responsibility of the authors and does not necessarily represent the official views of the National Institutes of Health.

The authors thank the members of the Massachusetts Higher Education Testing Group for motivating this study. Helpful conversations with Drs. Sandy Bogucki, Edward Kaplan, Stephanie Spangler, Madeline Wilson, and other members of the Yale University COVID-19 Public Health Committee shaped and refined our analysis, strategies and assumptions.

## Technical Appendix

### Model Description

We developed a dynamic, compartmental model using a modified “susceptible-exposed-infected-recovered” (or SEIR) framework. The model portrays the epidemiology and natural history of infection in a homogeneous population of at-risk individuals as a sequence of transitions, governed by difference equations, between different health states (or “compartments”). The model diagram (**Appendix Figure 1**, below) illustrates the modifications we made to the basic SEIR framework:

- Division of the “Infected” state into sub-compartments, to capture the important asymptomatic phase of COVID-19 disease and to distinguish it from the observable, symptomatic phase to which only some infected persons progress.
- Addition of the possibility of repeated screening with a test of imperfect sensitivity and specificity.
- Removal of infected individuals from the transmitting population based on either screening test findings or the development of COVID-defining symptoms.
- Removal (and return) of uninfected individuals from the transmitting population based on “false positive” screening test findings.
- Introduction of antibody protection, whether conferred via vaccination or recovery from prior infection, and the possibility of imperfect protection from infection, imperfect reduction in infectiousness, and imperfect reduction progression to more advanced COVID disease.
- Importation of additional new infections from exogenous sources (e.g., infections transmitted to students by university employees or members of the surrounding community. These are additional infections over and above those which would be predicted by the internal dynamics of the SEIR model.

We defined a total of 12 model states:

- U: Uninfected, susceptible
- E: Exposed, asymptomatic
- A: Infected, asymptomatic
- M: Infected, symptomatic, isolated
- TP: Infected, true positive, isolated
- FP: Uninfected, false positive, isolated
- UP: Uninfected, susceptible, antibody protected
- EP: Exposed, asymptomatic, antibody protected
- AP: Infected, asymptomatic, antibody protected
- TPP: Infected, true positive, antibody protected, isolated
- FPP: Uninfected, false positive, isolated, antibody protected
- D: Dead

Important groupings of these states include:

- <u>Active transmission and testing pool</u>. This pool is composed of individuals in states U, E, A, UP,

EP, and AP. All transmission of infection takes place between individuals in this pool. This is also the pool in which screening for infection takes place. Individuals in states U are susceptible to infection; so too are those in UP but at lower risk. Individuals in states A are able to transmit infection; so too are those in AP but at potentially lower rates. Persons in exposed states E and EP remain active in the transmission pool but have yet to incubate infection; consequently, they are unable to transmit, unable to be infected a second time, and unable to obtain a true-positive test result if screened.

- <u>Isolation pool.</u> The pool is composed of individuals in states M, FP, FPP, TP, and TPP.

Individuals in this pool are assumed to be isolated from the active transmission pool and from one another. We assume that proper infection control policies would prevent any nosocomial transmission; thus, infected and potentially infectious persons in states M, TP, and TPP do not transmit infection.

- <u>Antibody-protected pool.</u> Antibody protection is assumed to be conferred either via vaccination or via recovery from a prior infection. (We make no distinction between these two forms of protection.) The model permits the user to specify the degree to which antibody protection reduces susceptibility to infection from others, transmissibility of infection to others, and progression to symptoms in the event of a breakthrough infection.

#### Parameters

- β: rate at which infected individuals contact and infect susceptible individuals. This applies to transmission to persons in states A and AP.
- θ: incubation, the rates at which exposed individuals in states E and EP advance to asymptomatic, infectious compartments A and AP, respectively.
- σ: the symptom onset rate from states A and TP to state M. (We make the simplifying assumption that antibody protection shields asymptomatic, infected persons in state AP from progression to symptoms.)
- ρ: rate at which individuals in state i recover from disease. (We make the simplifying assumption that a single rate applies to states A, M, TP, AP, and TPP.)
- δ: the symptom-case fatality rate for individuals in state M.
- ε_i_: antibody effectiveness in reducing susceptibility to infection
- ε_t_: antibody effectiveness in reducing transmission

The model uses a cycle time of 8 hours. All rates are calculated per 8-hour cycle.

<u>Testing</u> is implemented in the model as a constant rate, governed by parameter τ for persons who are not antibody-protected and τp for persons who are not antibody-protected. This means that individuals are screened, at random, on average once every 1/τ cycles. This does not reflect the possibility of pulsed or scheduled screening at regular intervals. Note that there is a lag of one cycle between the time that a test is conducted and the time that a person receiving a positive test result is moved to isolation; the model is specifically designed to capture this delay. This captures the time to transport the sample to the lab, obtain the result, locate the individual, and effect the transfer to isolation.

- τ: rate at which non-antibody-protected individuals in the testing pool are screened for infection
- τ_p_: rate at which antibody-protected individuals in the testing pool are screened for infection
- Se: sensitivity of the screening test
- Sp: specificity of the screening test
- μ: rate at which false positives are returned from states FP and FPP to states U and UP, respectively.

Note: as currently implemented, the model permits parameter τ (the screening rate for non-antibody-protected individuals) to vary broadly between 0 and infinity. Parameter τ_p_ ( the screening rate for antibody-protected individuals) is only permitted to assume values 0 or τ.

<u>Exogenous shocks</u> makes it possible to consider the risk of superspreader events resulting from off-campus parties, infections transmitted to students by university employees or members of the surrounding community, or infections “imported” to campus from off-site sources. Users can choose to ignore this possibility or to explore its effects using two parameters:

- I(t): an indicator function which assumes value 1 if an exogenous shock takes place in cycle t and 0 otherwise;
- X: number of imported infections in a given exogenous shock (i.e., the magnitude of the shock), assuming that nobody is antibody-protected. (This value is adjusted to account for antibody protection in the susceptible population.)

#### Governing equations

For ease of presentation, we define the following intermediate calculations:

- **Active transmission pool at time t:**

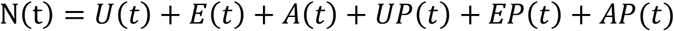

- **Endogenous Transmissions to persons in susceptible, non-antibody-protected state U at time t:**

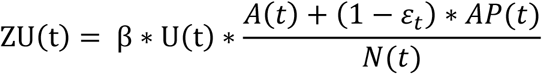

- **Endogenous Transmissions to persons in susceptible, antibody-protected state UP at time t:**

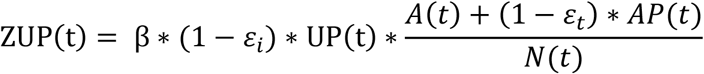

- **Vaccination-adjusted magnitude of exogenous shocks:**

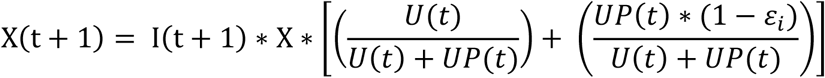

- **Exogenous Transmissions to persons in susceptible, non-antibody-protected state U:**

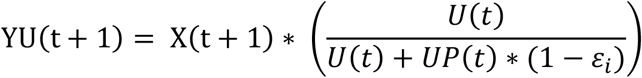

- **Exogenous Transmissions to persons in susceptible, antibody-protected state UP at time t:**

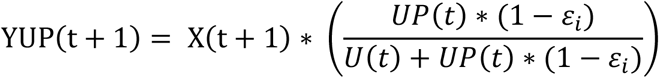

The equations governing transitions from one state to the next are:

- **Uninfected (t+1) = Uninfected (t) – False Positives - New Endogenous Transmissions – New Exogenous Transmissions + Returning False Positives**

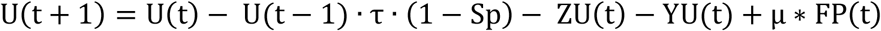

NOTE: The testing lag and some extreme parameter values create the possibility of depleting uninfected states U and UP. This equation (and the one for state UP) therefore includes a logic check (not shown) that prevents the population in states U and UP from falling below 0.

- **Exposed (t+1) = Exposed (t) - New Infections to A + New Endogenous Transmissions + New Exogenous Transmissions**

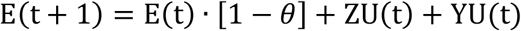
- **Asymptomatic (t+1) = Asymptomatic (t) – Symptom Onset – Recoveries – True Positives + New Infections**

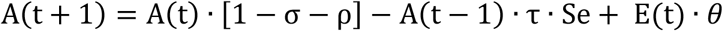
- **Mild/Moderate (t+1) = Mild/Moderate (t) – Recoveries - Deaths + Symptom Onset from A and TP**

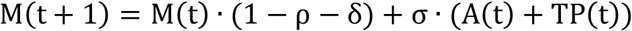
- **False Positives (t+1) = False Positives (t) – Returning FP+ New FPs**

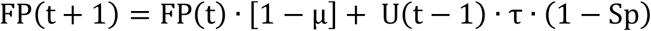
- **True Positives (t+1) = True Positives (t) – Symptoms – Recovery + New TPs**

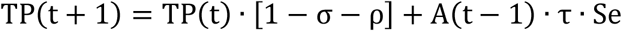
- **UninfectedProtected (t+1) = UninfectedProtected (t) – False Positives - New Endogenous Transmissions – New Exogenous Transmissions + Returning False Positives + Recoveries**

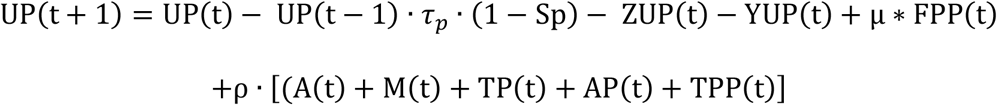
- **ExposedProtected (t+1) = ExposedProtected (t) - New Infections to AP + New Endogenous Transmissions + New Exogenous Transmissions**

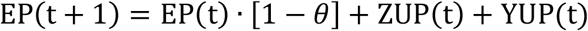
- **AsymptomaticProtected (t+1) = AsymptomaticProtected (t) – Recoveries – True Positives + New Infections**

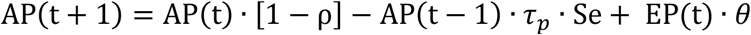
- **False PositivesProtected (t+1) = False PositivesProtected (t) – Returning FP+ New FPs**

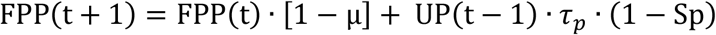
- **True PositivesProtected (t+1) = True PositivesProtected (t) – Symptoms – Recovery + New TPs**

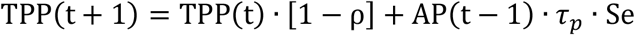
- **Deaths (t+1) = Deaths (t) + New Deaths**

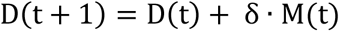
- **Total Population at time t:**

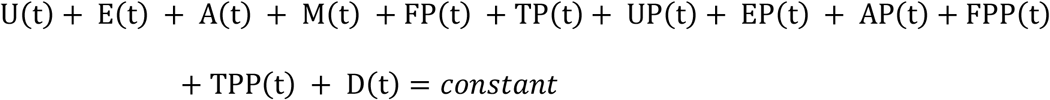

#### Initial conditions

We consider an active cohort of 5,000 individuals. Users may specify both the fraction of susceptible individuals who are antibody-protected (U(0) and UP(0)) and the number of unidentified, asymptomatic infections at time 0. Thus, states U(0), UP(0), and A(0) may assume positive values; all other state compartments are empty at time 0.

#### Parameter estimation

In this section, we describe how the parameters used in the governing equations are derived.

- **θ**: Individuals spend an average of 3 days in exposed states E and EP before progressing to Asymptomatic infection. This implies θ = (3 days)^-1^ = 0.333.
- **ρ**: We assume a time to recovery of 14 days, implying ρ =(14 days)^-1^ = 0.071.
- **σ:** We assume that 30% of persons with asymptomatic infection will advance to symptoms. Since this 30% value is determined by the formula σ/σ+ρ, this permits us to solve for σ = ρ(.3/.7) = 0.031.
- **δ**: We assume a symptom case fatality ratio of 0.5%. Since this value is determined by the formula δ/δ+ρ, this permits us to solve for δ = ρ(.005/.995) = 0.0000357.
- **β:** in the absence of antibody protection, the reproduction number R associated with this model is given by β / (σ+ρ). This permits us to solve for β = R * (σ+ρ) for the various values of R assumed in this analysis.
- **ε_t_ and ε_i_:** the model permits users to specify both εt a reduction in the rate of transmission (i.e., contagiousness) and εi a reduction in the rate of infection (i.e., susceptibility) in the presence of antibody protection. However, in the present analysis, parameter εt is conservatively set to 0, implying that antibodies have no effect in reducing contagiousness. Value of εI ranging from 50% to 100% are considered in sensitivity analysis.

**Appendix Figure 1.**
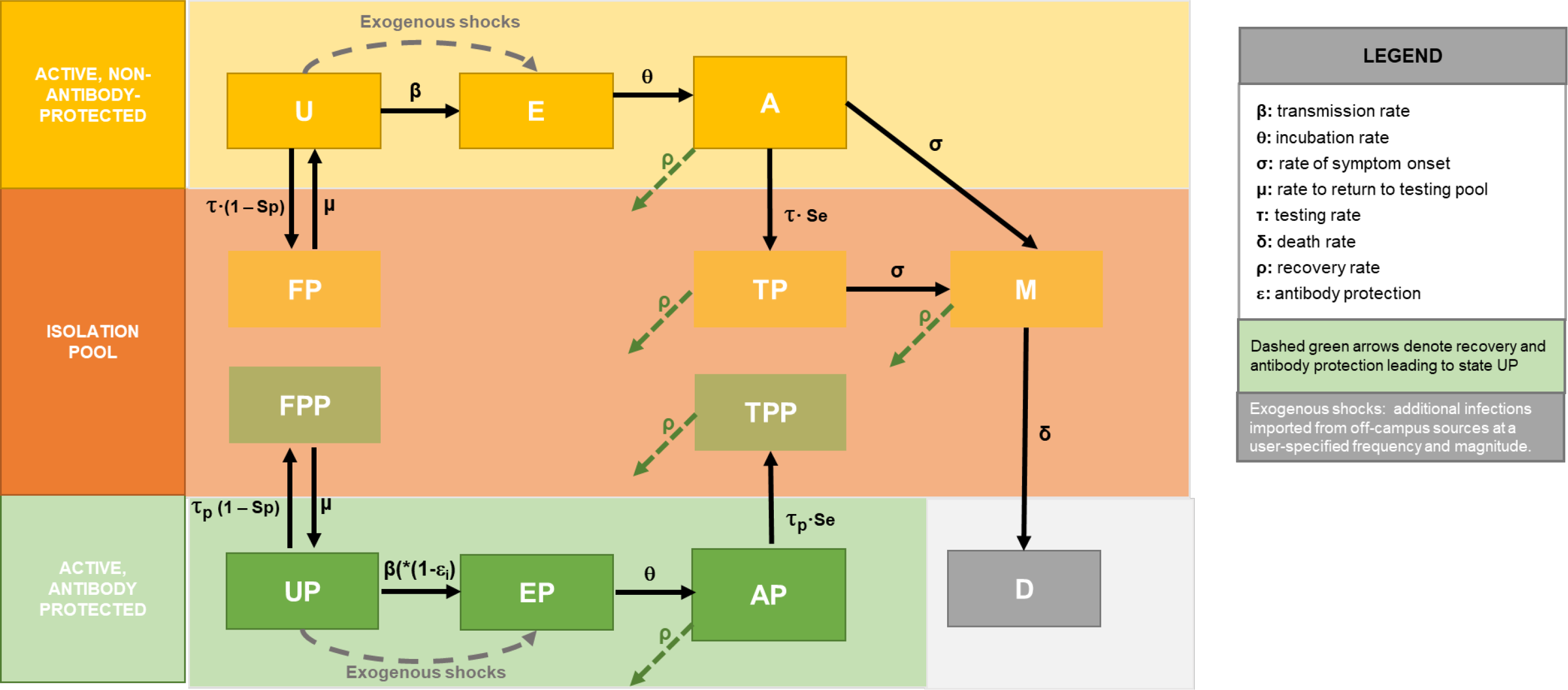

---

**Figure S1.**
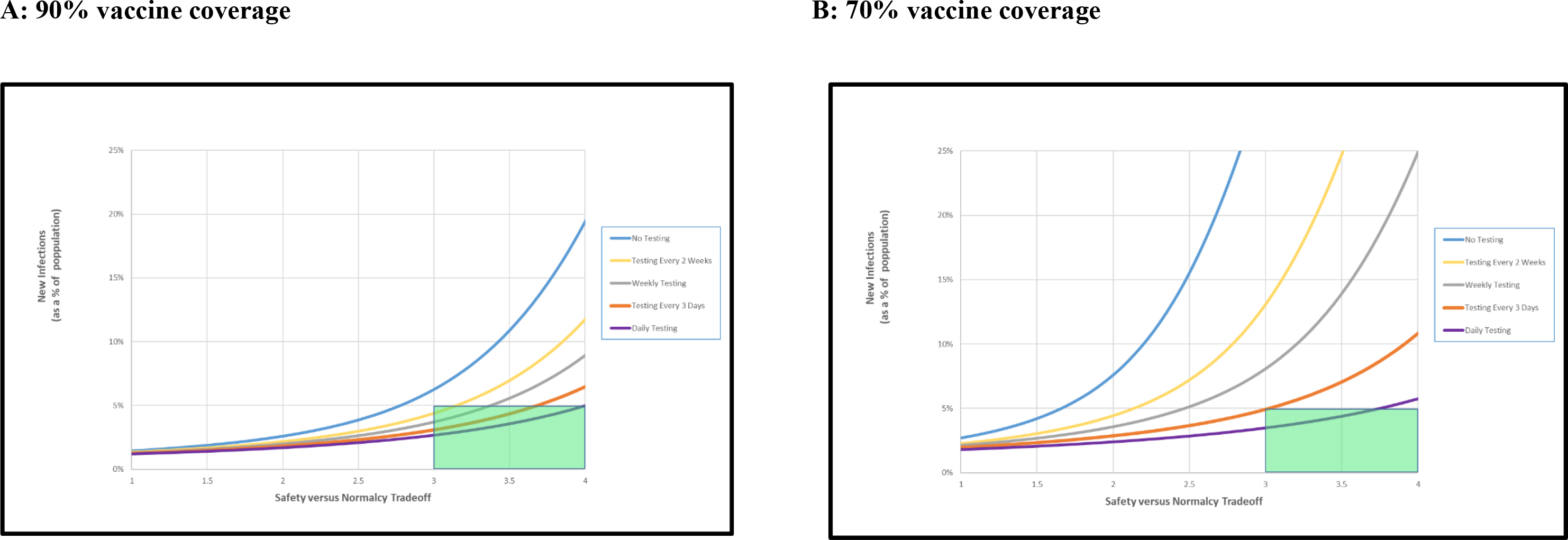
Sensitivity to vaccine effectiveness. The figure reports cumulative infections with vaccine effectiveness lowered to 80%. Asymptomatic testing is targeted to unvaccinated population. With 90% vaccination coverage (**Panel A**), infrequent testing (i.e., every 2 weeks) testing would be required to achieve the “return to normalcy” (Rt > 3, as defined previously) while holding cumulative infections below 5% of the campus population (green box). Daily testing would be required to achieve the same outcomes if the vaccination coverage rate was below 70% (**Panel B**).

**Figure S2.**
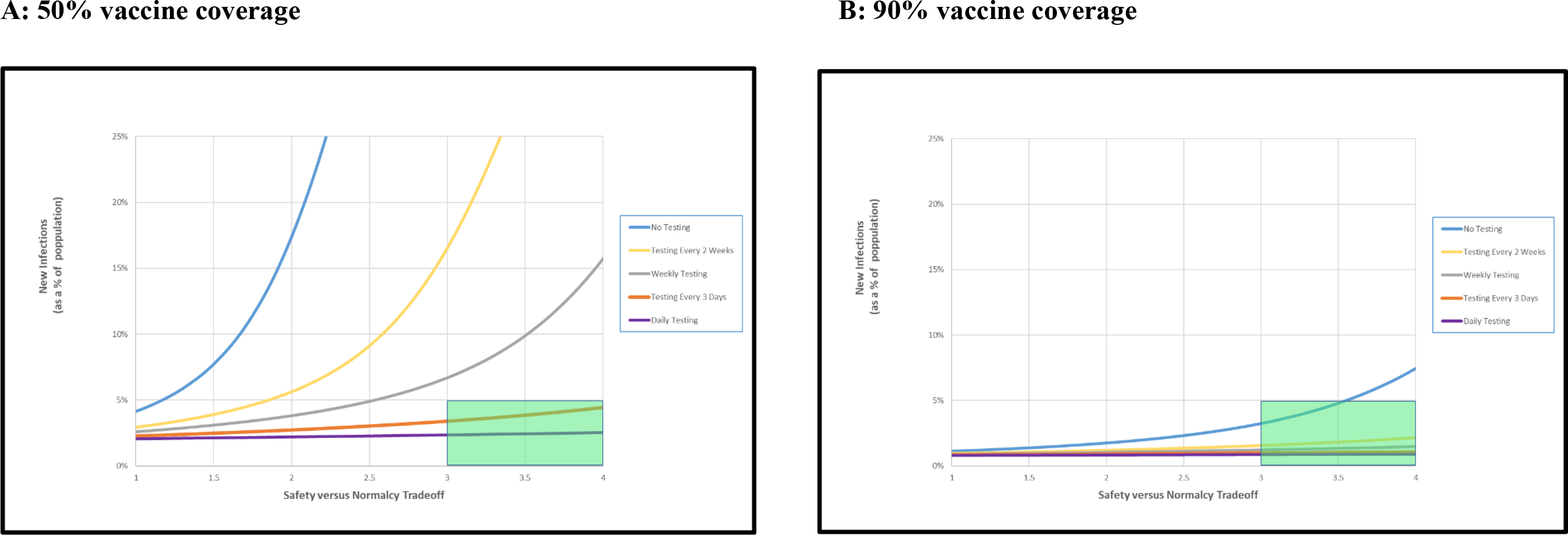
Sensitivity to test target expansion. The figure reports cumulative infections when routine, asymptomatic testing is extended to all members of the campus community, regardless of vaccination status. Vaccine effectiveness is at baseline value 85%. At colleges with low (50%) vaccination coverage (**Panel A**), the “return to normalcy” (Rt > 3, as defined previously) can be attained while holding cumulative infections below 5% of the campus population (green box) by testing the entire campus population once every 3 days. On campuses with vaccination coverage 90% (**Panel B**), expanding screening to the entire population would produce little discernible change from the base case.

